# Nurse-Led Telerehabilitation Intervention to Improve Stroke Efficacy: Protocol for a pilot randomized feasibility trial

**DOI:** 10.1101/2023.01.13.23284509

**Authors:** Stephanopoulos Kofi Junior Osei, Emanuella Adomako – Bempah, Adelaide Appiah Yeboah, Lawrence Akuamoah Owiredu, Lillian Akorfa Ohene

**Affiliations:** School of Nursing & Midwifery, University of Ghana, Accra Ghana; 37 Military Hospital, Accra Ghana; University of Ghana Hospital, Accra Ghana

**Keywords:** stroke, telerehabilitation, stroke nursing, telenursing

## Abstract

**Background:** The prevalence of stroke continues to rise in low-middle income countries. The continual rise in stroke cases and increasing prevalence on post-acute needs represent a crucial call for increased accessibility and utilization of rehabilitation services.

**Aim:** The primary objective of the study is to test the feasibility (recruitment, retention rates, cost and participant satisfaction) of a nurse-led telerehabilitation intervention in improving self-efficacy among stroke survivors. The findings of the trial are intended for use in a future larger study.

**Methods:** Participants would be recruited at the University of Ghana Hospital and randomized into an intervention group and a control group after being stratified based on age and type of stroke. Participants aged ≥ 18 years, diagnosed stroke at most 12months prior the recruitment and requiring moderate level of assistance would be considered for eligibility. Participants in the intervention group would undergo initial and continuous assessments for identification of problems and needs. Based on individual needs, participants would receive various nursing rehabilitation therapies in appropriate domains (cognitive, physical, emotional, and nursing education) combined with treatment as usual for 6 months. Participants in the controlled group would only receive treatment as usual (physiotherapy and follow-up with a medical officer). A follow-up evaluation would be conducted immediately, 30 days and 90 days after the intervention.

**Discussion:** Providing stroke rehabilitation services in low-resource settings presents a significant challenge due to limited infrastructure and a lack of trained healthcare professionals. The current study has the potential of contributing to the growing body of evidence on the impact of telerehabilitation services in mitigating these challenges in low-resource settings.

## Introduction

In low- and middle-income settings, stroke remains one of the major causes of disability and mortality [1-3].Patients who suffer a stroke may experience variable degrees of impairment and, in more serious situations, death. The aftermath of a stroke depends on several factors, including patient characteristics, emergency health-seeking behaviour, the nature of the stroke, and post-stroke sequelae[4-6]. It is crucial for stroke survivors with impairments to undergo rehabilitation as part of the recovery process [7].

According to Knect et al. [8], rehabilitation after stroke plays a critical role in improving functional outcomes, and mainly targets the recovery of motor and cognitive functions. Other studies have shown that providing emotional and psychological support for the patient and their caregivers during the rehabilitation process can greatly contribute to overall success [9,10].

Traditionally, stroke rehabilitation begins after the patient has been physiologically stabilized and is still on hospital admission [11]. This initial rehabilitation phase is primarily focused on improving physical functioning and facilitating the patient’s transition back to their home. This is followed by continuous rehabilitation, which typically takes place between 8 weeks and 6 months after the onset of the stroke. During continuous rehabilitation, patients make practical adjustments in their homes and try to navigate their natural environment with minimal assistance [12].

Despite the enormous impact of rehabilitation on recovery and life adjustment post-stroke, the delivery of equitable and affordable access to rehabilitation remains a challenge globally [13]. This challenge is more prominent in LMICs due to limited rehabilitation infrastructure and inadequate number of trained health workers.^14^ In Ghana, access to rehabilitation especially during the continued phase and post-hospital discharge is met with major challenges, including the high cost of available services, limited dedicated stroke rehabilitation services and units, socio-cultural barriers, long waiting times, poor education on rehabilitation and poor communication with healthcare providers [14,15]. As a result, persons recovering from stroke may have several unmet rehabilitative needs which remains unresolved, and adversely impact the recovery process [14].

These challenges exist partly due to the nature of conventional stroke rehabilitation during the continuous phase. Patients who are undergoing post-discharge rehabilitation mostly have to present themselves at the health facility, receive planned therapy and travel back to their homes [15]. This approach however is wrought with issues such as long travel distances and financial burden which often deter the stoke survivors from returning to use available services inf any. During the COVID-19 pandemic, such travels were virtually impossible in some settings creating a greater need to consider other innovative measures of continuing rehabilitative support even at a distance [16].

Telerehabilitation is becoming a highly considered approach to delivering stroke rehabilitation services. The approach has yielded positive outcomes, including improved stroke self-efficacy, motor functioning, and emotional well-being [17-19]. Telerehabilitation is projected to be a tremendous solution to the current challenges associated with rehabilitation accessibility. Additionally, stroke telerehabilitation has been found to be feasible and has received high acceptability among both patients and health workers in different countries, including Ghana [20,21].

With a limited number of stroke specialists in the country, stroke and community nurses in Ghana have to take a prominent role in supporting stroke rehabilitation [22]. As indicated earlier, the disabling effects of stroke put patients in a critical situation for increased rehabilitation support. Nurses are uniquely placed to support these patients after discharge. According to Kirvevold [12] nurses’ roles in post-discharge stroke rehabilitation includes but are not limited to coordinating treatment and care, providing assistance to survivors and family, providing informational support, consolation, monitoring condition progress, and supporting the resumption of roles and activities. These are all major roles that are crucial for improving patient outcomes and meeting the rehabilitation needs of stroke survivors and caregivers [23]. However, similar to conventional stroke rehabilitation, nurses’ involvement in stroke telerehabilitation is limited, particularly in LMICs [24]. Essentially, post-discharge stroke rehabilitation aligns with the mandate of nursing to promote recovery which places nurses at the core of the process. Despite this assertion, the critical role of nurses in post-discharge stroke rehabilitation especially in LMICs has not been well articulated in existing literature.

It is against this background that an innovative programme of care was developed. The current study seeks to assess the effects of a nurse-led stroke telerehabilitation intervention that facilitates the delivery of comprehensive nursing care to stroke survivors and caregivers in Ghana. The focus is to unearth the potential of telerehabilitation nursing in mitigating the numerous challenges affecting patients seeking stroke rehabilitation.

The study objective is to assess the preliminary feasibility and user satisfaction of a nurse-led holistic telerehabilitation among Ghanaian stroke survivors. The primary objectives of the trial are as follows:

1. Assess the retention of participants by estimating 1 month and 3-months follow-up rates
2. Assess the acceptability of nurse-led telerehabilitation intervention in terms of participant satisfaction
3. Examine participant experiences of nurse-led telerehabilitation intervention

A secondary objective of the trial is to measure whether a nurse-led telerehabilitation would improve self-efficacy of stroke survivors.

## Methods and analysis

### Study design

The trial is a parallel-group (intervention and control groups) randomized controlled study with 1:1 allocation to assess feasibility of a nurse-led telerehabilitation intervention administered remotely at the homes of Ghanaian stroke survivors. Post intervention process evaluation will employ a mixed method approach to exploring feasibility elements of the intervention.

### Study setting

Study participants would be recruited from the University of Ghana Hospital in Accra, Ghana. The facility has a dedicated stroke outpatient clinic for clients and a physiotherapy unit. We have already engaged with the chief nurse officer, lead physiotherapist, and head of the stroke clinic of the hospital, who are key to the success of the participant recruitment process.

### Eligibility criteria

Study participants will be considered for inclusion if they meet the following criteria:(1) is 18 years or above (2) diagnosed of a stroke with an onset of 1 week to 12 months prior the intervention (3) has FIM score of 55-100 (Level 3 moderate <u>assistance</u>) (4) total NIHSS score of 5 -15 (moderate stroke). However, study participants would be excluded subject to these criteria: (1) has no access to a smartphone (2) is unavailable for follow-up interviews (3) has a cognitive impairment (as documented in the patient’s medical records) that makes it difficult to comprehend the information provided and limits capacity to provide consent.

### Recruitment and screening

Participants would be recruited from the medical department and the stroke clinic of the University of Ghana Hospital. Patients at the study setting would be approached onsite and provided comprehensive background information about the study, including its risks and benefits during first encounter as part of inviting them to consider participating. If they opt to participate, eligibility clinical screening based on aforementioned criteria will be completed by the recruiting nurse. In this regard, the nurse will complete the FIM and NIHSS scores to ascertain eligibility. Informed consent would be obtained by researchers at this point (Fig 1). This informed consent has been approved by the Noguchi Institutional Review Board 032/21-22.

**Fig 1.** Enrolment, interventions and assessment schedule

### Intervention

Eligible participants would randomly be assigned to either the treatment or control group. Participants in control group would receive treatment as usual (TAU) which comprise of two weekly physiotherapy sessions and two sessions with a medical doctor.

Participants in the intervention group would receive comprehensive nursing neurorehabilitation support in addition to existing service at the hospital. The components of the intervention are guided by relevant literature, established guidelines [25-27] and have been reviewed by a panel of experts including two neuroscience nurses, two community health nurses, three neurologists, one stroke specialist, one physiotherapist and a stroke survivor. Patients would undergo assessment and receive various therapies in appropriate domains for 6months. A follow-up evaluation would be performed at the end, 30- and 90-days after the intervention (Fig. 2). The intervention would involve the following components;

**Fig 2.** Workflow

### Initial and ongoing patient assessment

Participants would undergo various assessments on motor function, cognitive capacity, anxiety and depressive levels and nutritional status utilizing evidenced-based assessment tools. The initial assessment would span in the first week of the intervention. Based on the problems and needs identified during the assessment, participants would receive a bundle of interventions in the domains of cognitive rehabilitation, physical rehabilitation, emotional support, and stroke education. These interventions would be provided twice a week in 1hr-to-1hr 30mins per session via video chats on WhatsApp. A participant in the intervention may have 192 sessions maximum or 48 sessions minimum in the span of the intervention.

### Cognitive rehabilitation domain

This domain is mainly focuses on supporting participants to establish a daily routine, improving memory and attention and mitigating spatial neglect [28]. We would guide participants in clearly structuring their daily activities based on their preferences and setting priorities [9]. Participants also be guided in visual scanning activities, attention training exercises and utilizing memory aids [29].

### Physical rehabilitation domain

The aim of physical rehabilitation will be the recovery of the patient’s motor functioning impacted by stroke [8]. Participants would undergo stroke-specific impairment assessment using the Fugl-Meyer Assessment (FMA) [30]. Based on identified needs the participant would undergo graded physical therapy including passive and activities range of motion exercises, constraint-induced exercises, and stress ball usage.

Additionally, participants and their caregivers would be educated on proper use of assistive devices for gait and mobility training.

### Emotional support

This domain is geared towards assessing and supporting the emotional status of participants and their caregivers. We would provide counselling sessions to help identify the extent of loss which greatly influences the emotional status and recovery of participants [31]. Additionally, study participants would be guided in recognizing their own emotions and identifying personalized but effective coping strategies [9].

### Stroke education

Education would be provided for participants and their relatives. The target of the information provided would focus on improving the participant’s understanding of the disease condition, modifiable risk factors of recurrence, pain management, preventing pressure ulcers, oral and catheter care, continence management, the importance of following professional dietary recommendations, and healthy nutrition in the recovery process [32].

See (Table 1) for details regarding the intervention presented based on the TiDieR checklist.

**Table 1:** Details of Intervention

| **Component of the intervention** | **What (procedures)** | **Who** | **How (mode of delivery)** | **Where (location of intervention)** | **When (schedule), How much, dose, intensity** | **Tailoring** |
| --- | --- | --- | --- | --- | --- | --- |
| Initial assessment | Comprehensive baseline patient assessment of the status of | Stroke nurses, and physiotherapist and | Via video call and on an | Patient's home | Only once as this serves as the baseline assessment. | All participants would undergo this |
|  | participants motor function, cognitive capacity, levels of anxiety and depression for the purpose of identification of participant needs | neuropsychologist as consultants. | individual basis |  |  | initial assessment for identification problem. |
| Ongoing assessment | Comprehensive ongoing patient assessment of the status of participants motor function, cognitive capacity, levels of anxiety and depression for the purpose of identifying patients progress and ascertaining possible deterioration | Stroke nurses | Via video call and on individual bases | Patient's home | Ongoing assessment would be provided as needed to fit participant needs changes and identifying progress | Ongoing assessment is participant specific and may not occur at similar frequency from one participant to the other |
| Cognitive rehabilitation domain | This domain focuses on guiding patients to establish a structured routine for every day, attention training exercises and learning the proper use of memory aids | Stroke nurses | Via video call and on individual bases | Patient's homes | Cognitive sessions would be held any other week participants with reported cognitive deficits that affect ADLs. Essentially, this group of participants would have 12 sessions on cognitive rehabilitation | Based on cognitive needs participants may be guided in using different strategies for either improving memory, or mitigating spatial neglect |
| Physical rehabilitation domain | Participants would undergo passive and/or active range of motion exercises, constraint-induced therapy and stress-ball | Stroke nurses, physiotherapist | Via video call and on individual bases | Patient's homes | Physical rehabilitations sessions would be held twice a week for participants with reported functional deficits that | Physical domains sessions are tailored to the functional needs of participants. Participants would be |
|  | usage.<br>Education on proper usage of assistive devices would also be provided |  |  |  | affect ADLs. Essentially, this group of participants would have 48 sessions on physical rehabilitation rehabilitation | grouped into those who can walk and stand with aids, those who cannot walk but stand with some gait issues and those who cannot stand at all. |
| Emotional support domain | Sessions would focus on understanding the extent of loss, recognizing emotions and mood changes and identifying personalised coping strategies for negative anxiety and depressive feelings that affect recovery. | Stroke nurses | Via video call and on individual bases | Patient's homes | Emotional support sessions would be held biweekly. Participants would have 12 sessions of emotional support. | Participants would have to the opportunity to identify individual coping strategies. |
| Stroke education domain | These sessions presents topics related to stroke as a conditions, modifiable risk factors, pain management, pressure ulcer prevention, nutrition and continence. | Stroke nurses | Via video call and on individual bases and group sessions | Patient's homes | These sessions would be held every three weeks during the intervention period. | Certain topics would be discussed during individual session to meet personal needs. |

### Outcomes and outcome measures

#### Primary outcomes

Based on the CONSORT checklist [33] the pilot study is primarily aimed at assessing the preliminary feasibility and acceptability of the intervention in the Ghanaian context.

##### Feasibility

Feasibility would be measured by retention rate, rate of adverse events and recording the details of resources used in implementing the intervention. Additionally, duration of sessions and cost of internet for the research team and study participants would be captured.

##### Participant satisfaction

This will include the use of a 12-item questionnaire to assess the level of satisfaction of participants on delivery of the intervention, time spent, quality of audio-visual channels involved and overall perceived benefits [34].

Participant experiences: Qualitative data of participant experiences would be captured in order to provide in-depth explanation of satisfaction levels. This would be guided by an interview probe designed for purpose.

#### Secondary outcome

##### Self-efficacy

The secondary and clinical outcome in focus would be Stroke Self-efficacy. The goal is assessed potential effectiveness of the intervention in improving self-efficacy among patients. This would be measured the Stroke Self Efficacy Questionnaire (SSEQ). The SSEQ is a 13-item scalar questionnaire that measures the stroke survivor’s ability and confidence in performing tasks under self-care, mobility and effective coping. The scale is also vital for assessing the functional performance of the study participants after the intervention [35,36]. This tool is most appropriate because it has been proven to have good face validity with a Cronbach Alpha of 0.90 suggesting good internal consistency and higher criterion validity when compared with other self-efficacy scales [37].

### Proposed sample size

We aim at recruiting 20 participants per group (total = 40 participants). The size however is guided by Julious’ rule of thumb for 12 participants per group in pilot trial [38]. The potential attrition rate estimated is 20% per group. During the follow-up phase, qualitative data on participant experiences would be captured from 15 conveniently sampled participants from the intervention group.

### Randomization, allocation and blinding

After eligible participants have provided informed consent and undergone baseline screening, they would be randomly assigned to the intervention group (Nurse-led rehabilitation support + Treatment as usual) and control group (only treatment as usual). A concealed block randomization (1:1) would be conducted by an independent researcher breaking sequentially numbered opaque envelopes, prepared by the said independent researcher using a random number generator [39]. Both participants and interventionists (nurses, physiotherapists, and neuropsychologists) would be unmasked during the study.

### Data collection, entry and management

Data would be gathered from participants before the intervention but after recruitment and after the intervention. Baseline data on patient history, socio-demographic characteristics, stroke self-efficacy levels, functional status, psychological function and cognitive capacity would be gathered from participants in both the intervention and control group. The second phase of data collection would take place immediately after the intervention capturing quantitative date on user satisfaction, qualitative data on experiences throughout the intervention only for participants in the intervention group and stroke-self efficacy levels for participants in both group. The third and final phase of data collection would be executed 4 weeks and 12 weeks after the intervention respectively capturing stroke efficacy levels in both groups. The data collection process would be undertaken by the investigators. After completing each phase of data collection, the quantitative data would be entered into a spreadsheet using Microsoft Excel. The data would be processed for outliers and missing inputs. Qualitative interviews would be audiotaped, audios would be saved in a PC folder and later transcribed for analysis. The entered information and saved audios would be protected by a password only accessible by the research team.

### Data analysis

Quantitative data analysis would be conducted using SPSS v26. The socio-demographic characteristics of participants would be summarized in frequencies and percentages. Continuous data gathered would be scrutinized for normality and necessary transformation would be applied if needed. Continuous variables would be summarized using means and SD. Mann-Whitney test or its parametric equivalent (if possible) would be employed to analyses the difference stroke self-efficacy levels in both groups at same points of follow-up evaluation. A repeated-measure analysis would be employed to ascertain changes in stroke self-efficacy level over the time. Results would be classified as significant if p <0.05 (two-tailed). All qualitative transcripts exported to NVivo version 10 would be analyzed by reading that data line by line and making noting down insights as meaning units while the reading is ongoing. The meaning units would be labeled with a code, this would be done repeatedly to capture all aspects of the content (data). The codes would then be categorized into sub-categories and broader categories for identification of themes [40].

### Possible risks and mitigation

There are minimal perceived risks to study participants involved in the intervention. However, all participants would undergo periodic risk assessments at their homes by the research investigators. Additionally, participants and their caregivers would be provided emergency dial lines and the contacts of the researchers in case of an adversity. All adverse events related to the intervention would be well-documented, reported to clinicians in charge and reviewed. Additionally, the research team has liaised with community health nurses to provide assessment and further recommendation for patients in their homes.

### Ethics and dissemination

The current study is targeted at determining the feasibility of a nurse-led telerehabilitation intervention for stroke patients in a low-resource setting. The study will illuminate the level of feasibility and effectiveness of this approach in settings similar to Ghana. Additionally, the study is crucial in further development and designing for similar interventions aimed at increasing accessibility to stroke rehabilitation using digitalisation. The study has ethical clearance from the Noguchi Memorial Institute of Research – IRB. Additionally, the trial has been registered in a public domain (PACTR202210685104862, Pan African Clinical Trial Registry). In the case of protocol modifications, both the IRB and trial registry would be informed using appropriate channels.

The findings of the study would be disseminated through peer-reviewed journals, nursing online and offline communities and conferences. Various key stakeholders including the Noguchi Memorial Institute of Research, the Ministry of Health Ghana and the University of Ghana Hospital (Legon) would be provided feedback and possibly recommendations based on the findings associated with the proposed study. Dataset at individual participant-level will not be shared indiscriminately. It would be made available on reasonable request.

## Discussion

Accessibility to stroke rehabilitation in low-resource settings remains a prevalent challenge due to limited infrastructure and inadequately trained healthcare professionals [13,14]. In Ghana, the challenges are exacerbated especially after hospital discharge by barriers including limited communication from health workers, socio-cultural barriers, inadequate rehabilitation education, high cost of available services and long waiting times.^15^

There is evidence that nurses – the largest health professional group in Ghana, are uniquely placed to support rehabilitation services and increase accessibility [12].

The current study is deemed to reveal the effects of a nurse-led telerehabilitation intervention on improving the self-efficacy of stroke survivors in a low-resource setting. The intervention is holistic and draws underpinning insights from relevant literature, a multidisciplinary team and patients.

Although the approach is therapy-based, the primary goal is to offer individualized and family-centred rehabilitation support coordinated by nurses. Langhorne, Bernhardt & Kwakkel^40^ asserts that multidisciplinary involvement, individualizing rehabilitation intervention and increasing intensity is crucial to the improvement of patient outcomes.

The study would also shed light on the feasibility of this emerging approach in delivering stroke rehabilitation services in a low-resource setting like Ghana. Additionally, the study would contribute to the existing literature on the potential impact of telehealth on healthcare delivery services in Ghana and other LMICs [41].

## Limitations

The study is being conducted on a feasibility scale and involves a small size which may not be sufficient to detect significant differences between study groups albeit the findings can offer useful insight into telerehabilitation services for stroke survivors. Additionally, the limited follow-up periods of the study limit the ability to explore long-term effects of the intervention.

## Conclusion

The implementation of a nurse-led telerehabilitation intervention for stroke survivors offers a potential contribution to mitigating the major challenges that threaten the accessibility of stroke rehabilitation services after acute management and post-hospital discharge.

## Data Availability

No datasets were generated or analysed during the current study. All relevant data from this study will be made available upon study completion.

## Abbreviations

IRB: Institutional Review Board
LMICs: Low and Middle Income Setting
PC: Personal Computer
TAU: Treatment
WHO: World Health Organization

## Author contributions

SKJO was responsible for formulating the idea of the study. SKJO, LAO (1), EAB and AAY were responsible for the literature review, identifying research gaps and the construction of the components of the study intervention. SKJO and AAY were responsible for the study design. LAO(2) was responsible for manuscript review and coordinating the study with SKJO.

## Acknowledgements

The authors would like to acknowledge the immense support offered by Miriam Iddrisu, Joyce Pwavra and Samuel Adjorlolo (PhD) all of the University of Ghana School of nursing and midwifery. We are also grateful to Dr Ram Hariharan, Dr Siva Nair, Nurse Hellen Oteng and Jonathan Bayuo (PhD) for their brilliant comments on this manuscript.

## Appendices

**S1 Fig**. Fig 1 Enrolment, interventions and assessment schedule

**S2 Fig**. Fig 2. Workflow Chart

**S3 Table**. Table 1: Details of Intervention

**S4 File. File 1** Ethical Clearance

**S5 File. File 2** SPIRIT Checklist

**S6 File**. **File 3** CONSORT Checklist

**S7 File. File 4** Protocol Approved by NIMR-IRB

